# Hydroxychloroquine and mortality risk of patients with COVID-19: a systematic review and meta-analysis of human comparative studies

**DOI:** 10.1101/2020.06.17.20133884

**Authors:** Thibault Fiolet, Anthony Guihur, Mathieu Rebeaud, Matthieu Mulot, Yahya Mahamat-Saleh

## Abstract

**Background:** Global COVID-19 deaths reached at least 400,000 fatalities. Hydroxychloroquine is an antimalarial drug that elicit immunomodulatory effects and had shown in vitro antiviral effects against SRAS-CoV-2. This drug divided opinion worldwide in the medical community but also in the press, the general public and in public health policies. The aim of this systematic review and this meta-analysis was to bring a new overview on this controversial drug and to assess whether hydroxychloroquine could reduce COVID-19 mortality risk in hospitalized patients.

**Methods and Findings:** Pubmed, Web of Science, Cochrane Library, MedRxiv and grey literature were searched until 10 June 2020. Only studies of COVID-19 patients treated with hydroxychloroquine (with or without azithromycin) compared with a comparative standard care group and with full-text articles in English were included. Studies reporting effect sizes as Odds Ratios, Hazard Ratio and Relative Risk for mortality risk and the number of deaths per groups were included. This meta-analysis was conducted following PRISMA guidelines and registered on PROSPERO (Registration number: CRD42020190801). Independent extraction has been performed by two independent reviewers. Effect sizes were pooled using a random-effects model.

The initial search leaded to 112 articles, from which 16 articles met our inclusion criteria. 15 studies were retained for association between hydroxychloroquine and COVID-19 survival including 15,081 patients (8,072 patients in the hydroxychloroquine arm and 7,009 patients in the standard care arm with respectively, 1,578 deaths and 1,423 deaths). 6 studies were retained for hydroxychloroquine with azithromycin. Hydroxychloroquine was not significantly associated with mortality risk (pooled Relative Risk RR=0.82 (95% Confidence Interval: 0.62-1.07, I^2^=82, P_heterogeneity_<0.01, n=15)) within hospitalized patients, nor in association with azithromycin (pooled Relative Risk RR=1.33 (95% CI: 0.92-1.92, I^2^=75%, P_heterogeneity_<0.01, n=6)), nor in the numerous subgroup analysis by study design, median age population, published studies (vs unpublished articles), level of bias risk. However, stratified analysis by continents, we found a significant decreased risk of mortality associated with hydroxychlroquine alone but not with azithromycin among European (RR= 0.62 (95%CI: 0.41-0.93, n=7)) and Asian studies (RR=0.36 (95%CI:0.18-0.73, n=1)), with heterogeneity detected across continent (P_heterogeneity between_=0.003). These finding should be interpreted with caution since several included studies had a low quality of evidence with a small sample size, a lack of adjustment on potential confounders or selection and intervention biases.

**Conclusion:** Our meta-analysis does not support the use of hydroxychloroquine with or without azithromycin to reduce COVID-19 mortality in hospitalized patients. It raises the question of the hydroxychloroquine use outside of clinical trial. Additional results from larger randomised controlled trials are needed

## Introduction

On December 31, 2019, World Health Organization (WHO) identified in Wuhan (China) an unknown pneumonia caused by a new coronavirus, SARS-CoV-2. This new coronavirus rapidly spread around the world and on the 11^th^ of March, the WHO declared it as a pandemic. By 17 June, 2020, WHO confirmed 8,006,427 cases and 436,899 deaths.

Recent publications identified the *in vitro* antiviral activity against SARS-CoV-2 of hydroxychloroquine (HCQ), an aminoquinoline like chloroquine. HCQ appeared as a potential treatment for COVID-19 patients at low costs(1). HCQ is also used as antimalarial drug, for rheumatoid arthritis and for lupus. This drug was widely advertised by international press and the United States President(2). Three *in vitro* studies tested HCQ on VeroE6 cells infected by SARS-CoV-2. This later suggested that HCQ decreased the viral replication with 50% inhibitory concentration (IC50) values of 2.2 µM (0.7 µg/mL) and 4.4 µM (1.4 µg/mL) in Maisonnasse *et al*. study, at 0.72 µM in *Yao* et al. study and between 4.51 – 12.96 µM for 50% maximal effective concentration (EC50) in Liu *et al*. study (1–3). Another study reported a synergistic effect of the HCQ with azithromycin (AZ) against SARS-CoV-2(6). The mechanism would be an acidification of the endosomes pH, and this pH modification would block the virus-endosome fusion (7).

Hydroxychloroquine was also tested in a study where macaques were infected by SARS-CoV-2 and received either a high dose of hydroxychloroquine (90 mg/kg on day 1 then 45 mg/kg) either a low HCQ dose (30 mg/kg on day 1 then 15 mg/kg) (3). Hydroxychloroquine did not improve the time to viral clearance. Another study in preprint also reported that there is no evidence of efficacy for the drug hydroxychloroquine (6.5 mg/kg) against infection with SARS-CoV-2 in hamsters or macaque models(8).

By June 17, about 132 trials have been referenced to test hydroxychloroquine for COVID-19 on ClinicalTrials.gov (9). Until today, most of the published studies on hydroxychloroquine with a comparative group (standard care) were observational and non-randomized with inconsistent results (10–16). This study is the first meta-analysis to pool adjusted relative risk and to include 16 studies. Previous meta-analysis on COVID-19 included a very limited number of studies and used unadjusted risk ratio (17–19). Thus, the aim of this meta-analysis was to provide a systematically quantitative assessment of the association between HCQ treatment (vs standard care) and COVID-19 survival risk among human trials and observational studies.

## Material and methods

### Data sources, search strategy

Research question was: does hydroxychloroquine treatment (vs standard care) have an effect (positive or negative) on survival of patients with COVID-19? A search was performed via PubMed and Web of Science and Cochrane Review until 10 June 2020 with this string search: (COVID-19 OR SRAS-CoV-2) AND (MORTALITY OR DEATH) AND (HYDROXYCHLOROQUINE OR HCQ) (Supplementary text S1). Given that the number of articles about hydroxychloroquine and COVID-19 is rapidly growing, we also manually searched additional reference on MedRxiv preprint server and on google scholar. The language was limited to English. This meta-analysis was conducted following PRISMA statements in Supplementary text S2. This study has been recorded on the international database of prospectively registered systematic reviews, PROSPERO (Registration number: CRD42020190801).

### Criteria for study selection

Inclusion criteria were 1) reports must contain original data with available risk estimates (Hazard Ration, Odds Ratios, Relative Risk and/or with data on the number of death in HCQ and control groups 2) all publication dates will be considered 3) publications in English language 4) comparative studies with a control group without hydroxychloroquine and 5) COVID-19 confirmed cases by RT-PCR. Reviews and meta-analysis, commentaries, *in vitro* and *in vivo* studies were excluded.

### Data extraction

Data extraction was performed by two investigators (Mr. T. Fiolet and Mr. Y. Mahamat-Saleh) who screened the titles and abstracts. Discrepancies were resolved by a third investigator (Dr. Anthony Guihur).

The following data were extracted from each study: study design, publication date, location, number of participants (total, in treatment and control groups, doses when available, effect size (Hazard Ratio, Odds Ratio or Relative Risk) and 95% confidence intervals for reported risk estimates. Hazard Ratio (HR) refers to the ratio of hazards in the intervention group divided by those occurring in the control group. Hazard represents the instantaneous event rate, which means the probability that an individual would experience an event (e.g. death) at a particular given point in time after the intervention, assuming that this individual has survived to that particular point of time without experiencing any event. In contrast, Relative Risk (RR) and Odds Ratio (OR) does not take account of the timing of each event. RR and OR are similar when the event (death) is rare. The most adjusted effect size reflecting the greatest control of potential confounders was extracted.

Three included studies did not report effect size for mortality risk (15,20,21). Thus we used the number of death per groups to calculate an unadjusted relative risk using *metabin* function in *meta* package in R Software (22). RR calculation is based on Cochrane Handbook for Systematic Reviews of Interventions formula 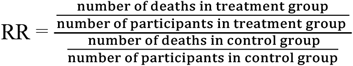 (23)

For all the other studies, reported adjusted OR, RR or HR were used. The quality of each study was assessed with ROBIN-I tool following Cochrane guidelines for non-randomized studies and with Rob2 for randomized studies (24,25).

### Outcome

The outcome is COVID-19 mortality.

### Statistical analysis

#### Effect of HCQ alone and HCQ + AZ

A primary meta-analysis was performed to assess the association between hydroxychloroquine alone (vs standard care) and risk of death. In a second time, the relationship between hydroxychloroquine associated with azithromycin and mortality was assessed. HRs, ORs and RRs were treated as equivalent measures of mortality risk. Pooled RRs were determined by using a random effect model with inverse variance weighting (DerSimonian-Laird method) (26). Significance was checked by Z-test (p<0.05 was considered as significant).

Heterogeneity was assessed by the Chi-square test and I^2^ test. 30%<I^2^<60% was interpreted as moderate heterogeneity and I^2^>60 as high heterogeneity. Funnel plot was constructed to assess the publication bias. Begg’s and Egger’s test were conducted to assess the publication bias (7,27). RR or HR and their 95% confidence interval were used to assessed mortality risk.

#### Subgroup analysis

Subgroup analyses were further conducted according to the quality assessment to explore the source of heterogeneity among observational studies. We performed stratified analyses by continents, the type of article (peer-reviewed vs unpublished), the use of an adjustment on confounding factors (studies with RR_unadjusted_ vs RR_adjusted_), the mean daily dose of hydroxychloroquine (continuous), the median population age across the studies (median age>63 years) and the level of bias risk identified with ROBIN-I (moderate/serious/critical) (24), the exclusion of studies with cancer and dialysis patients. Mean daily dose of hydroxychloroquine is a daily average between the loading dose and the maintenance doses. Additionally, influence analysis was conducted by omitting each study to find potential outliers (28). It is used to detect studies which influence the overall estimate of our meta-analysis the most, omitting one study at a time (leave-one-out method).

A two-sided p-value <0.05 was considered statistically significant. All analysis were conducted using R version 3.6.1 with *meta* package and *robvis* package (29).

## Results

### Literature Search

After searching Pubmed and Web of Science, 105 results were identified. 7 articles from Medrxiv/Google Scholar were added. After screening the title and the abstract, only 9 articles about hydroxychloroquine and COVID-19 were included. 144 articles were excluded for not meeting the inclusion criteria. 16 articles were included for further consideration including 14 observational studies and one non-randomized trial and one unpublished randomized controlled trial (RCT): 15 articles for HCQ (10–17,20,21,30–34) and 6 articles for HCQ+AZ (10,16,30,31,35,36). Flow chart is presented in Figure 1.

**Figure 1:**
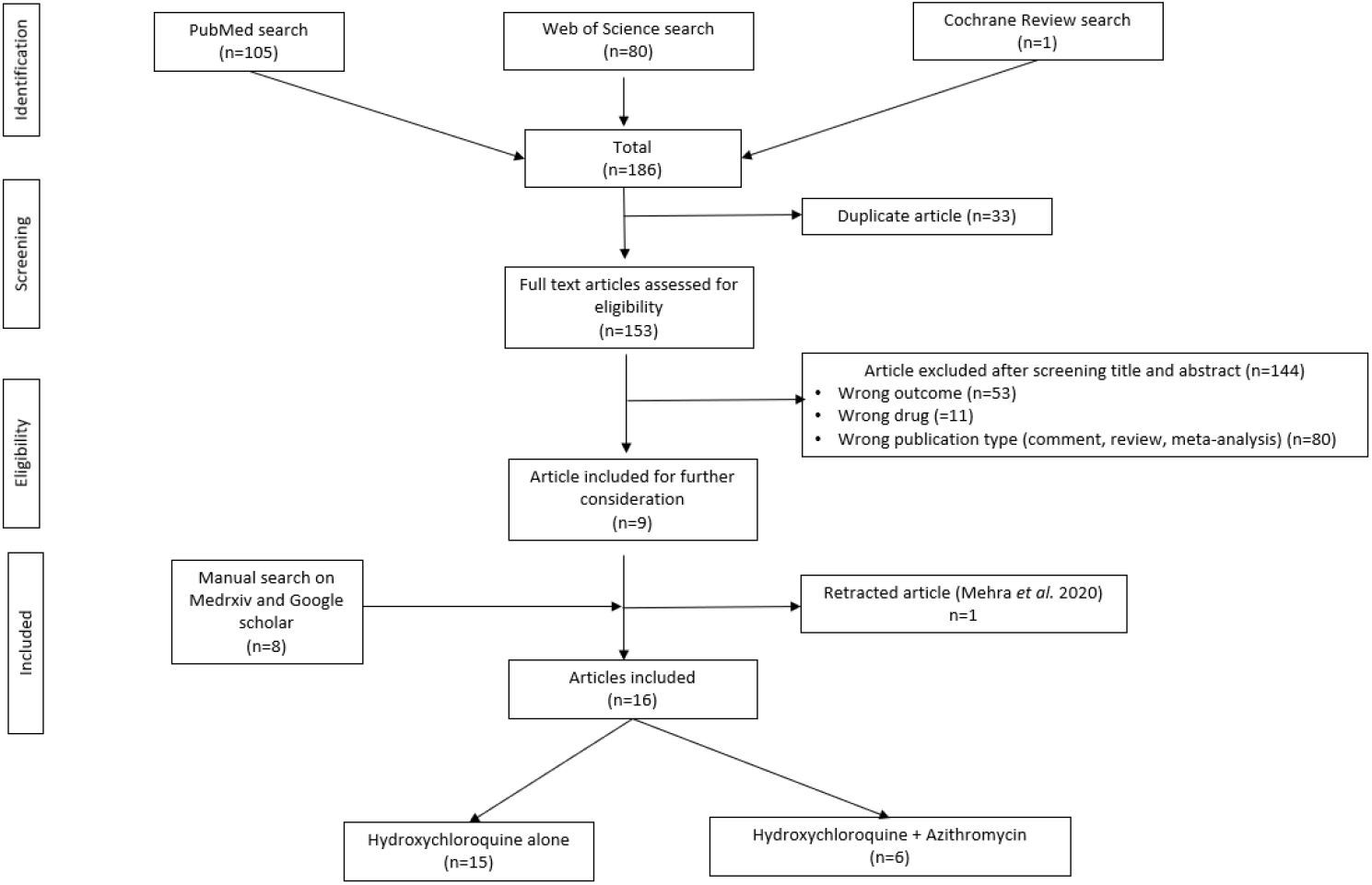
Flow diagram of study selection process.

### Study characteristics

This meta-analysis includes 8,072 patients in the hydroxychloroquine group and 7,009 patients in the standard care group with respectively 1,578 deaths and 1,423 deaths. Individual studies are described in Table 1. It appears that all the included studies were carried on hospitalized patients. No study meeting our inclusion criteria addressed the effect of HCQ on asymptomatic forms of COVID-19. Mean and median age of participants ranged from 53 to 72 across the studies. Studies were conducted in the USA (n=6) (13,16,20,30,31,36), in Spain (n=4) (14,15,33,35), in France (n=2) (11,21), in the UK (n=1)(37), in Italy (n=1) (32), in China (n=1) (12) and in 3 countries (USA, Canada and Spain)(10). 9 articles were published, and 4 articles were preprints. RECOVERY Trial data were reported by a press communication (34,37). Mean daily dose of hydroxychloroquine ranged from 333 mg/j to 945 mg/j.

**Table 1:** Characteristics of studies included in the meta-analysis for COVID-19 mortality

| First Author | Journal | Type of study | Country | Number of deaths |  | Number of participants |  | Treatment | Mean HCQ dose per day | Age<sup>a</sup> (years) | Patients | Study quality |
| --- | --- | --- | --- | --- | --- | --- | --- | --- | --- | --- | --- | --- |
|  |  |  |  | Control | HCQ | Control | HCQ |  |  |  |  |  |
| Alberici et al(32), 2020 | Kidney International | Observational, cohort | Italy | Not reported | Not reported | 22 | 72 | Not specified | NA | 72 (median)<br>IQ=62-79 | Hospitalized patients with haemodialysis | Critical |
| Ayerbe et al(15), 2020 | Journal of Thrombosis and Thrombolysis | Observational, cohort | Spain | 49 | 237 | 162 | 1857 | Not specified | NA | 67,57 (mean) | Hospitalized patients | Serious |
| Barbosa Joshua et al(20), 2020 | Unpublished | Observational, cohort | USA | 1 | 2 | 21 | 17 | 800 mg for 2 days then 200-400mg for 3-4 days | 600 | 62.7 (mean)<br>SD=15.1 | Hospitalized patients (mild/moderate symptoms) | Critical |
| Geleris et al(13), 2020 | NEJM | Observational, cohort | USA | 75 | 157 | 565 | 811 | 1200mg at day 1 then 400mg for 4 days | 560 | From <40 to >80 | Hospitalized patients (moderate/severe symptoms) | Moderate |
| Ip et al(31), 2020 | PrePrint | Observational, cohort | USA | 115 | 432 | 598 | 1914 | 800mg at day 1 then 400mg on day 2-5 (80%) | 400 | 64 (median)<br>IQ=52-76 | Hospitalized patients (44% moderate/severe symptoms) | Moderate |
| Kuderer et al(10), 2020 | The Lancet | Observational, cohort | USA<br>Canada<br>Spain | 41 | 11 | 486 | 89 | Not specified | NA | 66 (median)<br>IQ=57-76 | Hospitalized patients with who have a current or past diagnosis of cancer | Moderate |
| Magagnoli et al(30), 2020 | Clinical Advances | Observational, cohort | USA | 37 | 38 | 395 | 198 | Median HCQ dose: 400mg/day<br>Median HCQ+AZ dose: 422.2 mg/day | 400 | 70 (median)<br>IQ=60-75 | Hospitalized patients | Serious |
| Mahevas et al(11), 2020 | BMJ | Observational, cohort | France | 8 | 9 | 89 | 84 | 600mg/day | 600 | 60 (median)<br>IQ=52-68 | Hospitalized patients with covid-19 | Moderate |
|  |  |  |  |  |  |  |  |  |  |  | pneumonia who require oxygen: |  |
| Membrillo et al(14), 2020 | PréPrint | Observational, cohort | Spain | 21 | 27 | 43 | 123 | Loading dose of 800 mg + 400 mg in following days (ten days for moderate cases) | 440 | HCQ: 61.5<br>No HCQ: 68.7 (mean) | Hospitalized patients | Serious |
| Philippe Gautret et al(21), 2020 | International Journal of Antimicrobial Agents | Non-randomised controlled trial | France | 0 | 1 | 16 | 26 | A maintenance dose of 600 mg/day | 600 | 45,1 (mean)<br>SD=22 | Hospitalized patients (mild symptoms) | Critical |
| RECOVERY TRIAL | Unpublished | Randomized controlled trial | UK | 736 | 396 | 3132 | 1542 | A loading dose of 2400mg at day 1, then 800mg/day for 10 days | 945 | Not specified | Hospitalized patients | Not applicable |
| Rogado et al(35), 2020 | Clinical and Translational Oncology | Observational, cohort | Spain | Not reported | Not reported | 8 | 18 | Not specified | NA | 71 (median)<br>Range:34-90 | Hospitalized patients (64% severe cases) | Critical |
| Rosenberg et al(16), 2020 | JAMA | Observational, cohort | USA | 28 | 54 | 221 | 271 | 400mg then 200-400mg at 2nd prescription then 200-400mg at 3rd | 333 | 63 (median) | Hospitalized patients | Moderate |
| Sanchez-Alvarez et al(33), 2020 | Nefrología | Observational, cohort | Spain | 32 | 166 | 53 | 322 | Not specified | NA | 71<br>SD=15 | (85%) required hospital admission, 8% in intensive care units, with haemodialysis | Serious |
| Singh et al(36), 2020 | PrePrint | Observational, cohort | USA | 104 | 109 | 910 | 910 | Not specified | NA | 62<br>SD=17 | Hospitalized patients | Serious |
| Yu et al(12), | Science | Observational, | China | 238 | 9 | 502 | 48 | 400mg during | NA | 68 (median) | Critically ill | Serious |
| 2020 | China Life Sciences | cohort |  |  |  |  |  | 7-10 days |  | IQ: 59-77 | patients |  |

Table 1 (continued): Characteristics of studies included in the meta-analysis for COVID-19 mortality
| First Author | Effect size reported in each study<sup>b</sup> | Adjustments | Treatment | Control |
| --- | --- | --- | --- | --- |
| Alberici et al(32), 2020 | OR=0,44 [0,16-1,24] | Not adjusted | HCQ alone | Other antiviral and antibiotic were administered |
| Ayerbe et al(15), 2020 | RR<sub>calculated</sub>=0,422 [0,325-0,546] | Not adjusted | HCQ alone | Other antiviral and antibiotic were administered |
| Barbosa Joshua et al(20), 2020 | RR<sub>calculated</sub>=2,47 [0,24-24,98] | Not adjusted | HCQ alone | Supportive care |
| Geleris et al(13), 2020 | HR=1,04 [0,82-1,32] | inverse probability weighting from a propensity-score | HCQ alone | Standard care not specified |
| Ip et al(31), 2020 | HR=0.99 [0.8-1.22]<br>HR=0.98 [0.75-1.28] | Cox model adjusted on the propensity-score variable: gender, coronary disease, stroke, heart failure, arrhythmia, African American, COPD, , renal failure, rheumatologic disorder, inflammatory bowel disease, advanced liver disease, age, diabetes mellitus, insulin use prior to hospitalization, asthma, HIV/hepatitis, any cancer, and log ferritin | HCQ alone<br>HCQ+AZ | Group without drug |
| Kuderer et al(10), 2020<sup>c</sup> | OR=1,06 [0,51,2,2]<br>OR=2.93 [1.79-4.79] | Adjusted for age, sex, smoking status, and obesity | HCQ alone<br>HCQ+AZ | Treatment without AZ |
| Magagnoli et al(30), 2020 | HR=1.83 [1.16-2.89]<br>HR=1.31 [0.80-2.15] | Propensity score adjustment. All baseline covariates were included in the propensity score models (age, race, BMI, SpO2, breaths per minute, heart rate, T°, systolic blood pressure, ALT, AST, serum albumin, Total bilirubin, Creatinine, Erythrocytes, Haematocrit, Leukocytes, Lymphocytes, Platelets, Blood urea nitrogen, C-reactive protein | HCQ alone<br>HCQ+AZ | Standard care |
| Mahevas et al(11), 2020 | HR=1,2 [0,4,3,3] | Inverse probability of treatment weighting in Cox model. age, sex, comorbidities (presence of chronic respiratory insufficiency during oxygen treatment, or asthma, cystic fibrosis, or any chronic respiratory disease likely to result in decompensation during a viral infection; heart failure (New York Heart Association class III or IV); chronic kidney disease; liver cirrhosis with Child-Pugh class B or more; personal history of cardiovascular disease (hypertension, stroke, coronary artery disease, or cardiac surgery); insulin dependent diabetes mellitus, or diabetic microangiopathy or macroangiopathy; treatment with immunosuppressive drugs, including anticancer chemotherapy; uncontrolled HIV infection or HIV infection with CD4 cell counts <200/ $\mu$ L; or a haematological malignancy); body mass index ( $\geq 30$ or not); third trimester of pregnancy; treatment by angiotensin converting enzyme inhibitors or angiotensin receptor blockers<sup>13</sup>; time since symptom onset; and severity of condition at admission (percentage of lung affected: $\geq 50\%$ or not; presence of confusion; respiratory frequency; oxygen saturation without oxygen; oxygen flow; systolic blood pressure; and C reactive protein level). | HCQ alone | Standard care |
| Membrillo et al(14), 2020 | OR=0,07 [0,012,0,402] | Adjusted on variables with $p < 0,25$ in univariate analysis | HCQ alone | Standard care + other antivirals, immunomodulators, anti-inflammatory drugs |
| Philippe Gautret et al(21), 2020 | RR<sub>calculated</sub> =3,41 [0,1505,77,45] | Not adjusted | HCQ alone | Group without HCQ |
| RECOVERY TRIAL | HR=1,1 [0,98,1,26] | Adjustment not precised | HCQ alone | Standard care |
| Rogado et al(35), 2020 | OR=0,02 [0,01,0,73] | Adjusted by median age, histology, staging, cancer treatment received and hypertension | HCQ+AZ | Group without HCQ |
| Rosenberg et al(16), 2020 | HR=1,08 [0,63,1,85]<br>HR=1.35 [0.76-2.4] | Multiple adjustments on potential confounders (age>65, sex, hospital, comorbidities, respiratory capacities) | HCQ alone<br>HCQ+AZ | Group without drug |
| Sanchez-Alvarez et al(33), 2020 | OR=0,471 [0,28,0,792] | No information about adjustments in logistic regression | HCQ alone | Standard care + other antivirals |
| Singh et al(36), 2020 | HR=0,95 [0,74,1,23]<br>HR=1.19 [0.89-1.60] | Creation of groups based on propensity score matching for age, gender, race, confounding comorbidities | HCQ alone<br>HCQ+AZ | Group without HCQ |
| Yu et al(12), 2020 | HR=0,36 [0,18,0,75] | Adjustment: respiratory rate, shortness of breath, alanine aminotransferase (when $p < 0,01$ in univariate Cox model) | HCQ alone | Standard care |

IQ=Interquartile range, SD=Standard Deviation, HCQ=Hydroxychloroquine, AZ=Azithromycine, NA=Not available
<sup>a</sup>Some studies did not report mean or median age
<sup>b</sup>HR and OR are the most adjusted effect size reported in each study. Some studies did not report effect size. RR<sub>calculated</sub> were calculated using the number of death in the treatment and the control groups

### Study quality

Risk of bias was assessed with ROBIN-I for non-randomised studies (n=14) and Rob2 was not applicable for RECOVERY RCT because data were not available (Figure S1). Details on the assessment of studies quality are provided in Fig S2. Among the non-randomized studies, the majority of these observational studies had a high or critical risk of bias (10 out of 16) (12,14,15,20,21,30,32,33,35,36). Five articles had a moderate risk of bias(10,11,13,16,31). Some studies did not report adjusted effect sizes to control confusion and selection bias (15,20,21,32,33,35). Studies quality was lowered by the lack of information about the assignment of treatment, the time between start of follow-up and start of intervention), some unbalanced co-intervention with other antiviral and antibiotic drugs.

### Hydroxychloroquine and mortality

The pooled RR for COVID-19 mortality was 0.82 (95% CI: 0.62-1.07, I^2^=82, P_heterogeneity_<0.01, n=15) (Figure 2) indicating no significant association between hydroxychloroquine and COVID-19 survival or increased mortality. There was significant high heterogeneity across the included studies (I^2^ =83%, p<0.01). Egger’s test (p= 0.42) and Begg’s test (P=0.88) were not significant for asymmetry of the funnel plot indicating that there is not a major publication bias (Figure S3). In our separated analysis by study design, we found a positive but not significant association between hydroxychloroquine alone and mortality among interventional studies (RR: 1.10, 95%CI: 0.97-1.25, I^2^=0%, P_heterogeneity within_=0.5, n=2); however an inverse but not significant association was found among observational studies (RR: 0.78, 95%CI: 0.58-1.05, I^2^=82%, P_heterogeneity within_ <0.01), with heterogeneity observed across the study design (P_heterogeneity between_ = 0.03).

**Figure 2:**
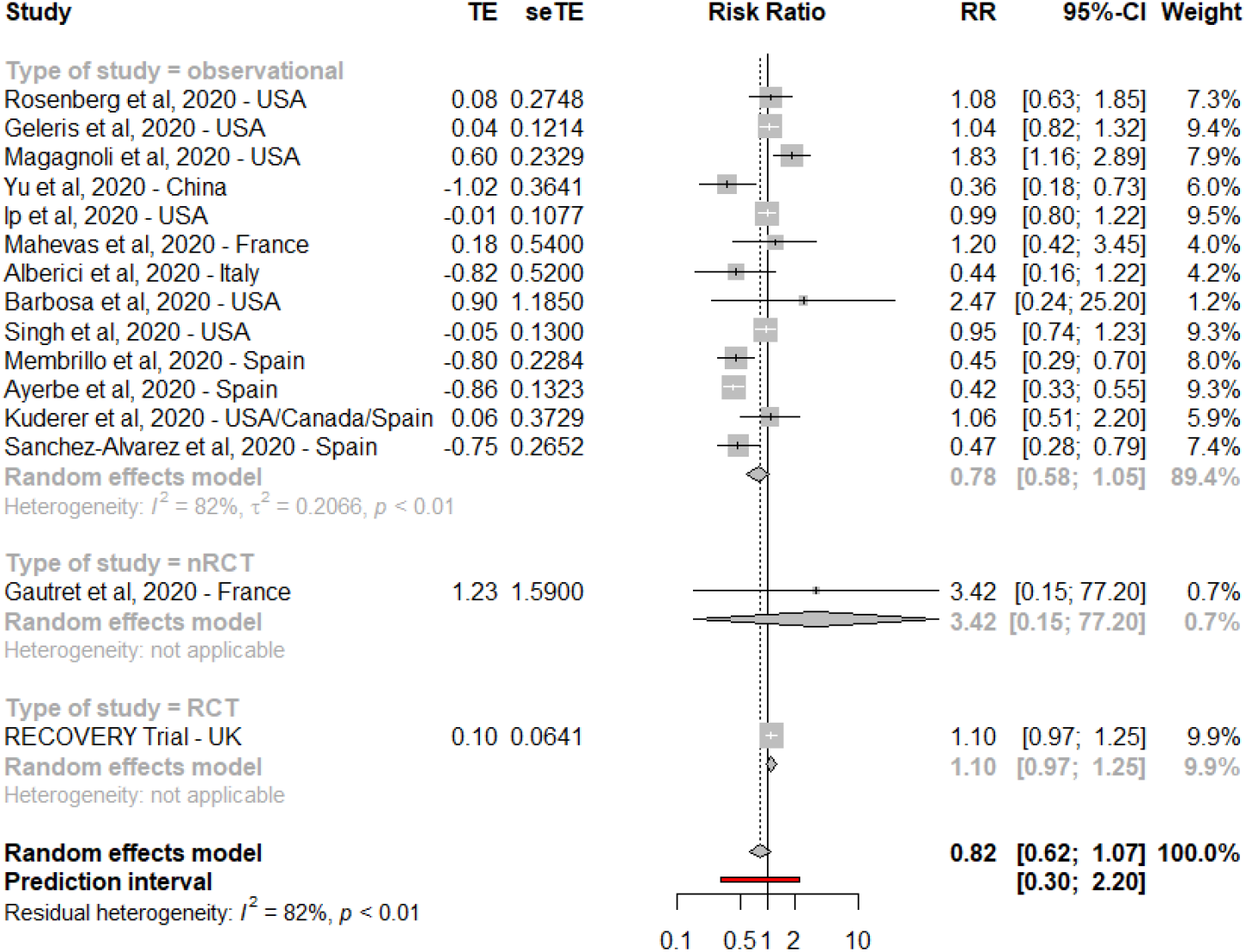
Meta-analysis showing association between hydroxychloroquine alone and COVID-19 mortality. RCT=Randomised Controled Trial. nRCT=non-Randomised Controled Trial TE=Estimated treatment effect. seTE=Standard error of treatment estimate. RR=Risk ratio. RR were not adjusted for Alberici et al, Ayerbe et al, Barbosa et al, Sanchez-Alvarez et al and Gautret et al. 95%CI= 95% Confidence Interval

### Subgroup analysis for hydroxychloroquine alone

Subgroup analysis among all studies (observational and interventional studies) per study design, type of article (peer-reviewed vs unpublished), risk estimated, age, the exclusion of cancer/haemodialysis patients identified a non-significant association in each subgroup (Table 2).

**Table 2.** Subgroup analysis for the associations between HCQ alone or HCQ associated with AZI and mortality risk of patients with COVID-19 (observational and interventional studies) N: number of studies. NA: Not applicable for a single study

|  | N | Pooled Relative Risk | Heterogeneity |  |  |
| --- | --- | --- | --- | --- | --- |
|  |  |  | I<sup>2</sup> (%) | P<sub>within</sub> | P<sub>between</sub> |
| **HCQ alone** |  |  |  |  |  |
| All Studies | 15 | 0.82 [0.62-1.07] | 82% | <0.01 |  |
| Study Design |  |  |  |  |  |
| Observational | 13 | 0.78 [0.58-1.05] | 82% | <0.01 | 0.03 |
| Interventional | 2 | 1.10 [0.97-1.25] | 0% | 0.48 |  |
| Type of article |  |  |  |  |  |
| Peer-reviewed | 10 | 0.78 [0.53-1.16] | 83% | <0.01 | 0.63 |
| Unpublished | 5 | 0.88 [0.63-1.24] | 74% | <0.01 |  |
| Adjusted estimate |  |  |  |  |  |
| Yes | 9 | 0.91 [0.67-1.24] | 70% | <0.01 | <0.0001 |
| No | 5 | 0.44 [0.35-0.56] | 0% | 0.41 |  |
| Missing | 1 | 1.10 [0.97-1.25] | Not applicable | Not applicable |  |
| Risk estimated |  |  |  |  |  |
| Reported in the paper | 12 | 0.86 [0.66-1.12] | 73% | <0.01 | 0.9 |
| Calculated | 3 | 0.91 [0.21-3.93] | 48% | 0.14 |  |
| Risk of bias |  |  |  |  |  |
| Moderate | 5 | 1.02 [0.88-1.18] | 0% | 0.9 | 0.19 |
| Serious | 6 | 0.63 [0.38-1.04] | 89% | <0.01 |  |
| Critical | 3 | 0.97 [0.23-4.07] | 31% | 0.23 |  |
| Continents |  |  |  |  |  |
| America | 6 | 1.05 [0.93-1.19] | 30% | 0.2 | 0.003 |
| Asia | 1 | 0.36 [0.18-0.73] | NA | NA |  |
| Europe | 7 | 0.62 [0.41-0.93] | 90% | <0.01 |  |
| Multiple | 1 | 1.06 [0.51-2.20] | NA | NA |  |
| Mean daily dose |  |  |  |  |  |
| Not specified | 6 | 0.58 [0.39-0.85] | 80% | <0.01 | 0.007 |
| <500 mg/d | 4 | 0.97 [0.55-1.69] | 84% | <0.01 |  |
| >500 mg/d | 5 | 1.09 [0.98-1.22] | 0% | 0.88 |  |
| Age |  |  |  |  |  |
| 63 years or less | 7 | 0.90 [0.65-1.26] | 53% | <0.05 | 0.1 |
| 64 years or more | 7 | 0.69 [0.43-1.10] | 88% | <0.01 |  |
| Not specified | 1 | 1.10 [0.97-1.07] | NA | NA |  |
| Cancer or haemodialysis patient based-population |  |  |  |  |  |
| No | 12 | 0.87 [0.64-1.18] | 84% | <0.01 | 0.26 |
| Yes | 3 | 0.61 [0.35-1.06] | 43% | 0.17 |  |
| Influence analysis (exclusion of Yu et al, Magagnoli et al, Membrillo et al, Ayerbe et al) | 11 | 1.00 [0.90-1.12] | 29% | 0.17 |  |
| **<u>HCQ+AZI</u>** |  |  |  |  |  |
| All Studies | 6 | 1.33 [0.91-1.91] | 75% | <0.01 |  |
| Study Design |  |  |  |  |  |
| Observational | 6 | 1.33 [0.91-1.91] | 75% | <0.01 |  |
| Interventional | 0 |  |  |  |  |
| Type of article |  |  |  |  |  |
| Peer-reviewed | 4 | 1.55 [0.86-2.80] | 76% | <0.01 | 0.2 |
| Unpublished | 2 | 1.07 [0.88-1.30] | 0% | 0.35 |  |
| Adjusted estimate |  |  |  |  |  |
| Yes | 6 | 1.33 [0.91-1.91] | 75% | <0.01 |  |
| No | 0 |  |  |  |  |
| Risk estimated |  |  |  |  |  |
| Reported in the paper | 6 | 1.33 [0.91-1.91] | 75% | <0.01 |  |
| Calculated | 0 |  |  |  |  |
| Risk of bias |  |  |  |  |  |
| Moderate | 3 | 1.54 [0.80-2.95] | 86% | <0.01 | 0.06 |
| Serious | 2 | 1.22 [0.95-1.57] | 0% | 0.7 |  |
| Critical | 1 | 0.02 [0.00-0.73] | NA | NA |  |
| Continents |  |  |  |  |  |
| America | 3 | 1.10 [0.91-1.32] | 0% | 0.48 | 0.0009 |
| Asia | 0 |  |  |  |  |
| Europe | 2 | 0.24 [0.00-13.43] | 80% | 0.02 |  |
| Multiple | 1 | 2.93 [1.79-4.79] | NA | NA |  |
| Mean daily dose |  |  |  |  |  |
| Not specified | 3 | 0.75 [0.08-7.21] | 87% | <0.01 | 0.7 |
| <500 mg/d | 3 | 1.10 [0.87-1.38] | 0% | 0.43 |  |
| >500 mg/d | 0 |  |  |  |  |
| Age |  |  |  |  |  |
| 63 years or less | 2 | 1.22 [0.94-1.58] | 0% | 0.69 | 0.9 |
| 64 years or more | 4 | 1.30 [0.62-2.71] | 85% | <0.01 |  |
| Cancer or haemodialysis patient based-population |  |  |  |  |  |
| No | 6 | 1.33 [0.91-1.91] | 75% | <0.01 |  |
| Yes | 0 |  |  |  |  |

Test for subgroup differences (observational vs nRCT vs RCT) was not significant (P=0.09) suggesting no differences in the overall effect according to the design of the studies. The pooled RR for observational studies was 0.78 (95%CI: 0.58-1.05, I^2^=82%, P_heterogeneity within_ <0.01, n=13) and RR was 3.42 (95%CI: 0.15-77.20, n=1) for non-randomized controlled trial and 1.10 (95%CI: 0.97-1.25, n=1) for the RECOVERY randomized controlled trial (Figure 2).

After stratification by the level of bias from ROBIN-I evaluation, the association between hydroxychloroquine and COVID-19 mortality remained non-significant. The broadness of 95% CI and heterogeneity increased with the risk of bias: moderate risk of bias (RR=1.02 [0.88-1.18], I^2^=0, P_heterogeneity within_ =0.9, n=5), serious risk of bias (RR=0.63, 95% CI: (0.38-1.04, I^2^=89%, P_heterogeneity within_ <0.01, n=6)) and critical risk of bias (RR=0.97, 95% CI: (0.23-4.07, I^2^=31%, P_heterogeneity within_ =0.2, n=3)) (Figure S4).

In our stratified analysis by continents (Figure S5), interestingly, we found a significant decreased risk of mortality with HCQ alone among Asian (RR_Asia_=0.36, 95%CI: 0.18-0.73, n=1) and European studies (RR_Europe_=0.62 (95%CI: 0.41-0.93, I^2^=90%, P_heterogeneity within_ <0.01, n=7)) but there was no significant association among American studies, with heterogeneity detected across continent (P_heterogeneity between_=0.003).

Furthermore, we found no association between HCQ alone and mortality by HCQ daily mean dose. The pooled RR was 1.09 (95%CI: 0.98-1.22, I^2^=0%, P_heterogeneity within_=0.9, n=5), for studies with >500mg, (RR=0.97 (95%CI: 0.55-1.69, I^2^=84%, P_heterogeneity within_<0.01, n=4) for HCQ dose<500 mg and (RR=0.58 (95%CI: 0.39-0.85, I^2^=80%, P_heterogeneity within_<0.01, n=6) for an unspecified dose of HCQ, with heterogeneity detected across HCQ dose categories (P_heterogeneity between_=0.007).

In our stratified analysis by studies which reported adjusted effect sizes (vs non-adjusted), the pooled RR for adjusted estimates was RR=0.91 (95%CI: 0.67-1.24, I^2^=70%, P_heterigeneity within_<0.01, n=9) and for non-adjusted estimates RR=0.44 (95%CI: 0.35-0.56, I^2^=0%, P_heterigeneity within_<0.41, n=5), suggesting differences in the overall effect according to the presence of adjustment on potential confounders.

Influence analysis showed that Yu et al, Membrillo et al, Ayerbe et al, Magagnoli et al are influent studies (Figure S7). Removing these studies make heterogeneity decrease at I^2^=0% but the results remained non-significant (RR=1.00 (95% CI: 0.0-1.13, I^2^=29%, n=11) (Table 2).

All the results remained similar after exclusion of the two interventional studies (Table S1).

### Hydroxychloroquine with azithromycin and mortality

The pooled RR for COVID-19 mortality was 1.33 (95% CI: 0.91-1.921, n=6) (Figure 3) indicating no significant association between hydroxychloroquine with azithromycin and survival. There was significant high heterogeneity across the included studies (I^2^ =75%, p<0.01). Egger’s test (p= 0.9) and Begg’s test (p=0.6) were not significant but the asymmetry in the funnel plot indicates that there could be a publication bias. However, the number of included studies is small.

**Figure 3:**
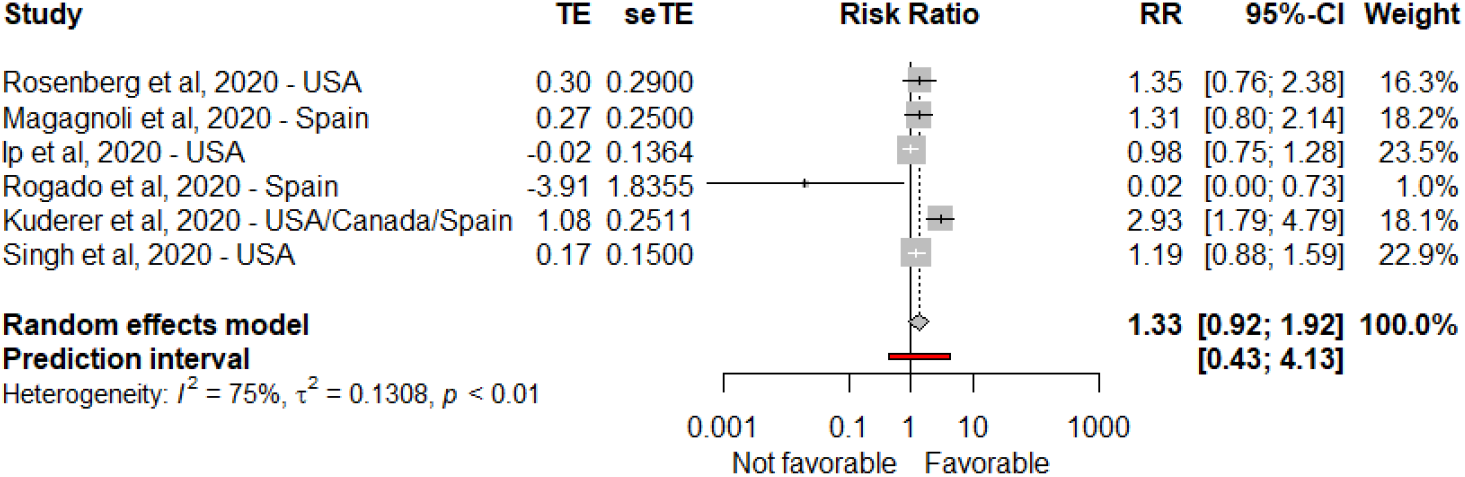
Meta-analysis showing association between hydroxychloroquine with azithromycin and COVID-19 mortality. TE=Estimated treatment effect. seTE=Standard error of treatment estimate. RR=Risk ratio. 95%CI= 95% Confidence Interval

### Subgroup analysis for hydroxychloroquine with azithromycin

In all the subgroup analysis (type of article, effect size, risk of bias, continent, mean daily dose, age, exclusion of cancer and haemodialysis patients, influence analysis), no significant association between hydroxychloroquine with azithromycin and mortality was found (Table 2). Nevertheless, in our stratified analysis by continents, we found no significant association with COVID-19 survival risk among American studies (RR=1.10, 95%CI: 0.91-1.32, I^2^=0%, P_heterogeneity within_=0.48, n=3) and European studies (RR=0.24 (95%CI: 0.00-13.43, I^2^=80%, P_heterogeneity within_ <0.02, n=2)) but there was a significant increased risk of mortality in the multiple countries (RR=2.93, 95%CI: 1.79-4.79, n=1), with heterogeneity detected across continent (P_heterogeneity between_=0.0009).

## Discussion

This meta-analysis summarized the results of 14 observational studies, 1 non-randomised study and 1 unpublished randomised controlled trial on hydroxychloroquine with or without azithromycin and COVID-19 survival (Table 1). The results indicated that hydroxychloroquine with or without azithromycin is ineffective to reduce COVID-19 mortality risk in hospitalized patients (Figure 2 and 3). Eight observational studies reported no advantage for hydroxychloroquine (10,11,13,16,20,21,31,32). One US Veterans study identified an increased risk of death(30). Three Spanish and one Chinese studies reported a protective effect (12,14,15,33) but this benefit on survival was not replicated in two RCT, especially RECOVERY Trial which is one of the largest study. Our meta-analysis reported a high heterogeneity. The use of an adjusted effect size to control confusion bias, the daily HCQ dose, the risk of bias and the localisation of the study (by continents) may explain one part of the heterogeneity observed according to our subgroup analysis.

Subgroup analysis revealed that there was a decreased risk of death among 6 European non-randomised studies, one observation Asian study and for studies which did not specify the treatment dose. However, five (14,15,21,32,33) of these European studies have a serious or critical risk of bias (Figure S1). This significant relationship could be explained by a high risk of confusion bias since these articles did not reported adjusted effect size. These studies also have several biases, such as a selection bias Gautret et al, control and treatment groups did not come from the same hospital. In 3 Spanish studies (14,15,33), there was no information when treatment were administrated and when the follow-up began which may lead to a bias in selection. Studies with an adjusted HR in figure S5 and with a higher quality reported a non-significant higher RR than the other studies. In this meta-analysis, the majority of the included studies had a high or critical risk of bias (10 out of 16) (Figure S1 and S2). Most of them do not always report the concomitant use of antiviral or antibacterial drugs. In our subgroup analysis by study design, we found inconsistent results with a positive but not significant association between hydroxychloroquine alone and mortality among interventional studies and an inverse but not significant association among observational studies (Table 2). Heterogeneity between these subgroups was observed across the study design. However, these findings are limited by the very low number of interventional studies.

Two Chinese randomised controlled trial reported no death in both treatment and control group (38,39) and thus their results were not included in our meta-analysis. A previous review on 8 studies (11–14,20,30,39,40) on COVID-19 concluded that the level of evidence for hydroxychloroquine effect is very weak(41). A preprint meta-analysis, using routinely collected records from clinical practice in Germany, Spain, the UK, Japan, and the USA, compared the use of HCQ vs salfasalazine (42). This study observed an increased risk of 30-day cardiovascular mortality (HR=2.19 [1.22-3.94]) but there was no standard care comparative group. Some previous meta-analyses were also conducted on hydroxychloroquine and various health endpoints including mortality. However these studies did not report all the published and unpublished literature, including a very limiting number of studies: from 3 articles(17,18) to 6 articles(19). These previous meta-analyses did not perform subgroup and sensitivity analysis to test the effect of pooling RCT and observational study, neither studying the source of heterogeneity. They used unadjusted risk ratio (calculated with the number of events in each group) whereas in our meta-analysis, we used adjusted relative risk (43) and we did sensitivity analysis on the adjustment of effect size. Statistical adjustments for key prognostic variables allow to limit confusion bias, especially in observational studies which are not randomised. Our meta-analysis confirmed the partial preliminary results of these meta-analyses about the absence of effect for HCQ on survival.

Our study has several strengths. To our knowledge, this is the first meta-analysis using adjusted relative risk and including numerous subgroup analysis (by continent, population age, effect size, risk of bias, published articles, mean daily dose of hydroxychloroquine, exclusion of cancer and haemodialysis patients) which found stable and consistent results. This study informs clinicians and patients regarding the efficiency of HCQ in treating COVID-19. We included several unpublished papers to minimize the publication bias. Our subgroup analysis by published studies (vs unpublished studies) identified that the inclusion of preprints did not change the results. Exclusion of grey literature (unpublished studies, with limited distribution) could lead to an exaggeration of the intervention effect by 15% (44). There is limited evidence to identify whether grey studies have a poorer methodological quality than published studies(45). Mortality is a reliable endpoint across studies. Limitations come from the studies which do not report adjusted effect size when mortality was not the primary endpoint. Confounding bias is high in these articles (mainly for the preprints). This meta-analysis was based on aggregated data, without access to original patient data. Most of studies are observational which do not allow to identify a causal association. This meta-analysis did not include results from the European DISCOVERY trial and the WHO SOLIDARITY trial (46). To finish, some of the included studies had very low quality of evidence (missing data, small sample size, confusion bias, bias in classification of intervention and selection bias) but the exclusion of these articles did not change the results.

Few peer-reviewed studies with a comparative group analysed some other endpoints such as virological clearance, clinical improvement and arrhythmia risks. A recent randomized controlled trial with 821 asymptomatic participants in contact with a COVID-19 confirmed case, concluded hydroxychloroquine was not efficient to prevent illness in a prophylactic way (47). However, this trial had a limitation: only 16 participants had a confirmed positive RT-PCR test. A small French non-randomised trial identified a higher proportion of negative RT-PCR tests in the HCQ group (21) but two other RCT did not find any difference between the HCQ and standard care groups for clinical improvement (38,39).

Several studies raised concerns about an increase of the QTc interval with HCQ use in an intensive care unit (48) and hospitalized patients (11,49). However, this side effect was not found in Tang et al. RCT. Several national health organisations (US FDA Food and Drug Administration(50), French Agency for the Safety of Health Products ANSM (51), European Medecine Agency EMA(52)) raised concerns about using this unapproved drug for COVID-19. ANSM et US FDA removed the authorization for its use outside of clinical trials. The Indian Council of Medical Research took an opposite position and recommend chemoprophylaxis with hydroxychloroquine for asymptomatic cases (53). In an open label, randomised controlled trial with hydroxychloroquine in patient with mild and moderate symptoms, no death were reported (38). Finally, in the comparative peer-reviewed studies, a clear conclusion on hydroxychloroquine is not possible due to the small sample size, the lack of well-performed randomised controlled trials (mainly non-randomised and retrospective studies) and inconsistent results. Many preprints without comparative group and without randomization bring confusion in this highly politicised topic. There is a gap between the speed of clinical research and the expectation of a clear solution to treat COVID-19 patients. Indeed, producing robust clinical trials is necessarily time-consuming. Results from large RCT are needed to shut down the controversy.

## Conclusion

In conclusion, there is no strong evidence supporting a benefice for hydroxychloroquine with or without azithromycin to improve survival of COVID-19 hospitalized patients. Conversely, there is no strong evidence supporting an increased mortality associated with HCQ or HCQ + AZ intake.

## Data Availability

The authors confirm that the data supporting the findings of this study are available within the article included in the meta-analysis

## Abbreviations

HCQ: Hydroxychloroquine
AZ: Azithromycin
RR: Relative Risk
HR: Hazard Ratio
OR: Odds Ratio
US FDA: US Food and Drug Administration
EMA: European Medicine Agency
CI: Confidence Interval

## List of figures and tables

Figure 1: Flow diagram of study selection process

Figure 2: Meta-analysis showing association between hydroxychloroquine alone and COVID-19 mortality (observational vs intervention)

Figure 3: Meta-analysis showing association between hydroxychloroquine with azithromycin and COVID-19 mortality.

Table 1: Characteristics of studies included in the meta-analysis for COVID-19 mortality

Table 2: Subgroup analysis for the associations between HCQ alone or HCQ+AZI and mortality risk of patients with COVID-19 (observational and interventional studies)

## Supplementary tables and figures

Supplementary tables and figures

S1. Full electronic search strategy

**Figure S1:**
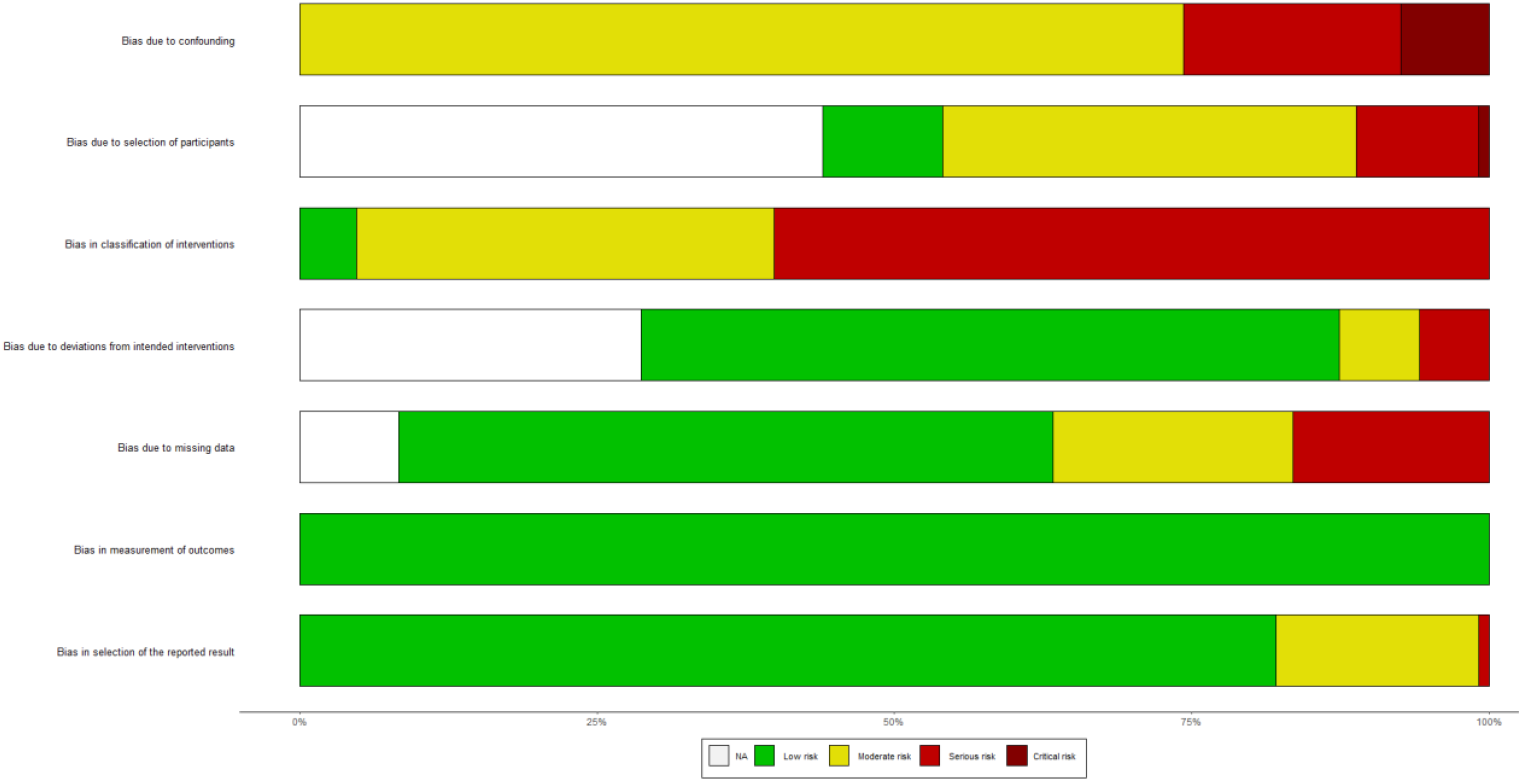
Summary of risk of bias analysis for non-randomised studies (ROBIN-I)

**Figure S2:**
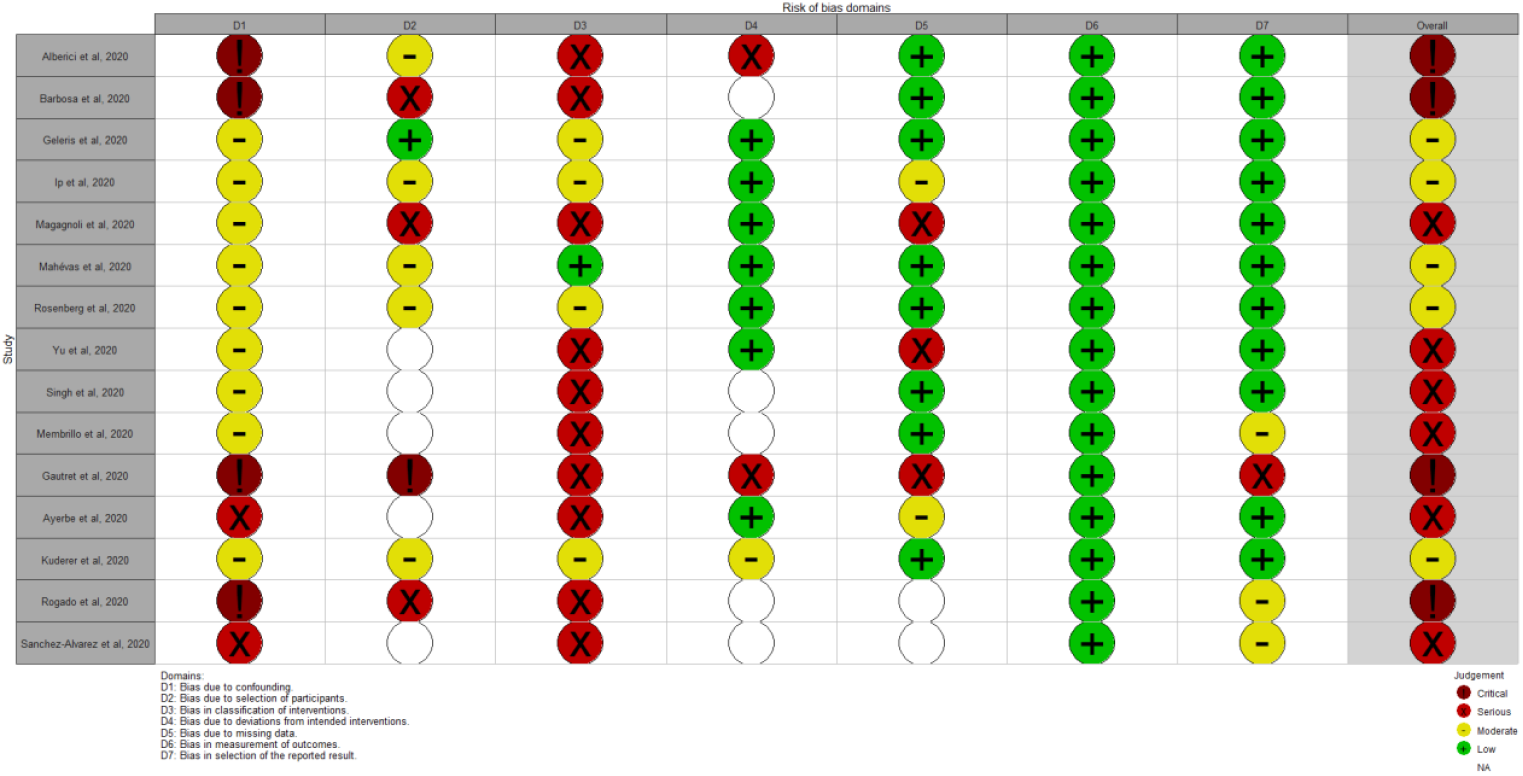
Assessment of quality of studies using ROBIN-I for non-randomised studies.

**Figure S3:**
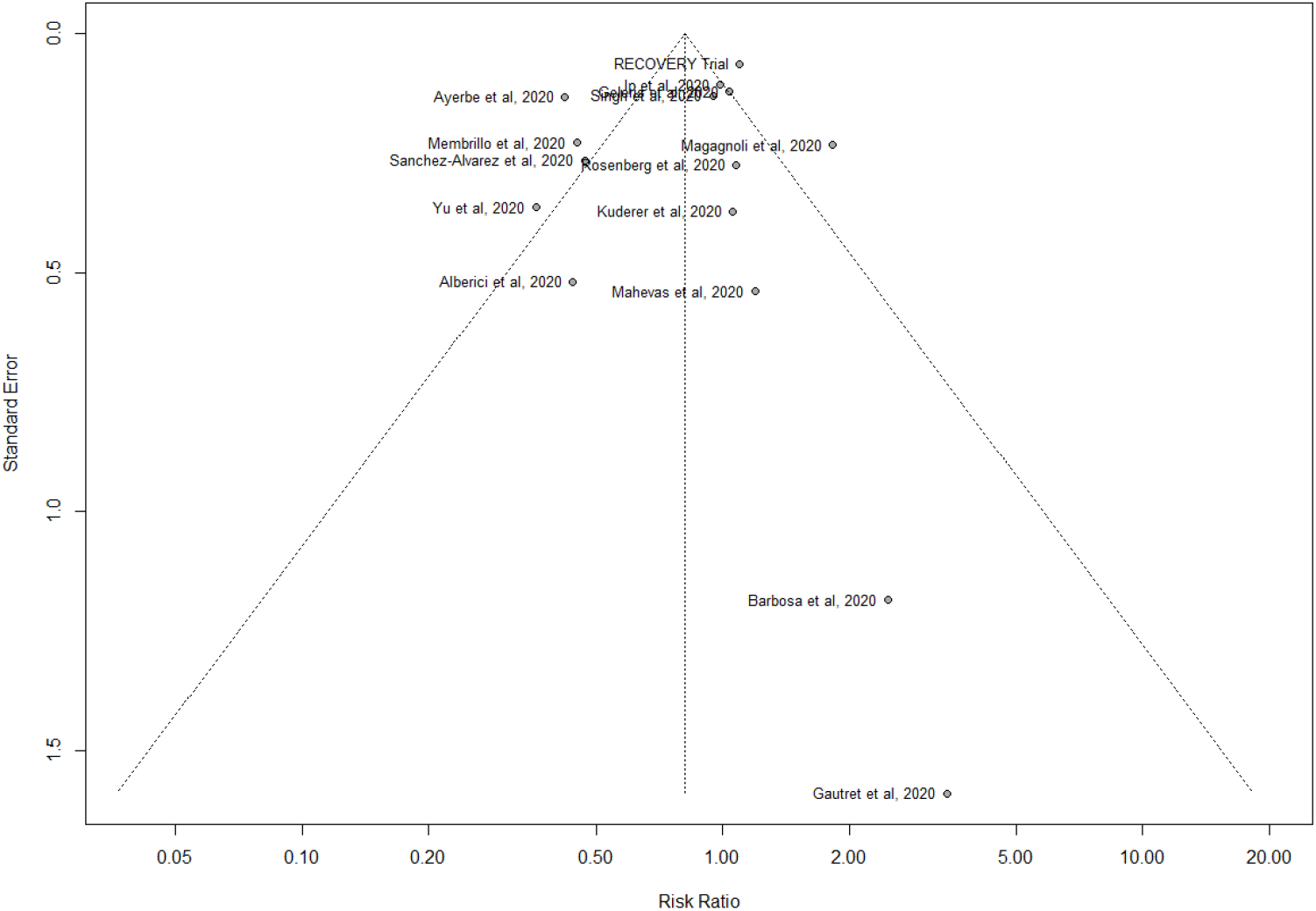
Funnel plot for hydroxychloroquine alone and COVID-19 mortality risk

**Figure S4:**
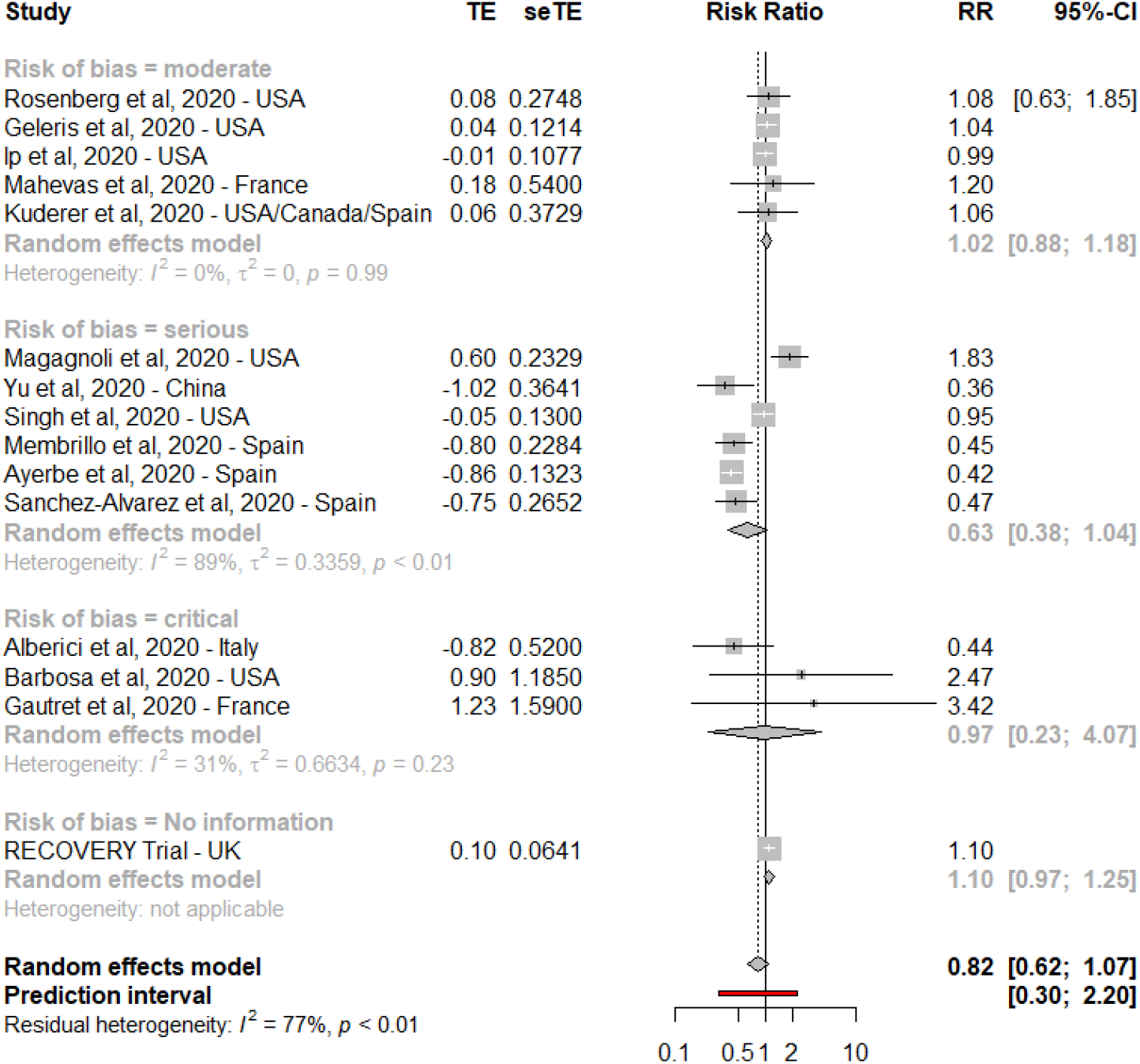
Forest plot for hydroxychloroquine alone and COVID-19 mortality risk, subgroup analysis per risk of bias

**Figure S5:**
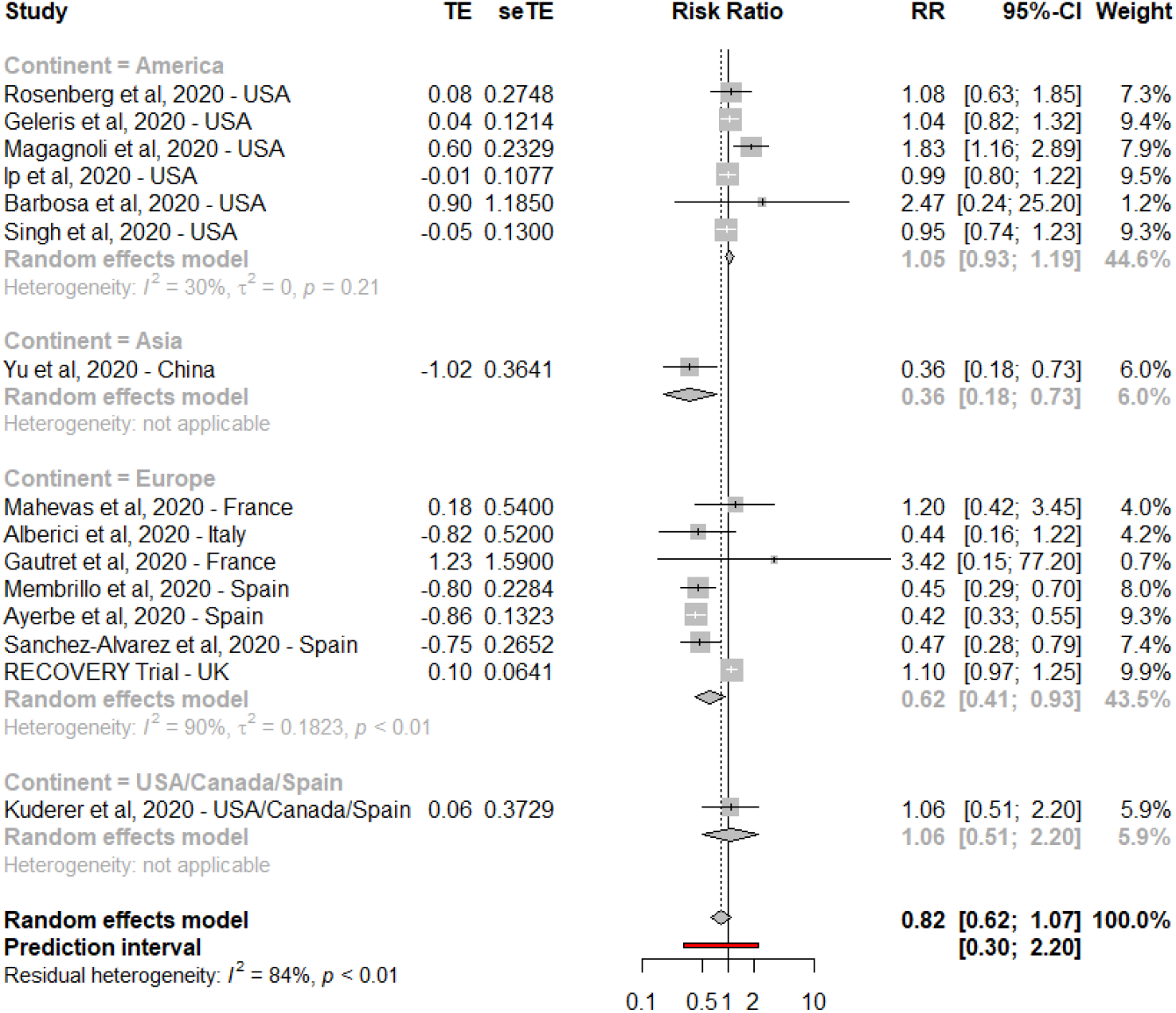
Forest plot for hydroxychloroquine alone and COVID-19 mortality risk, subgroup analysis per continent

**Figure S6:**
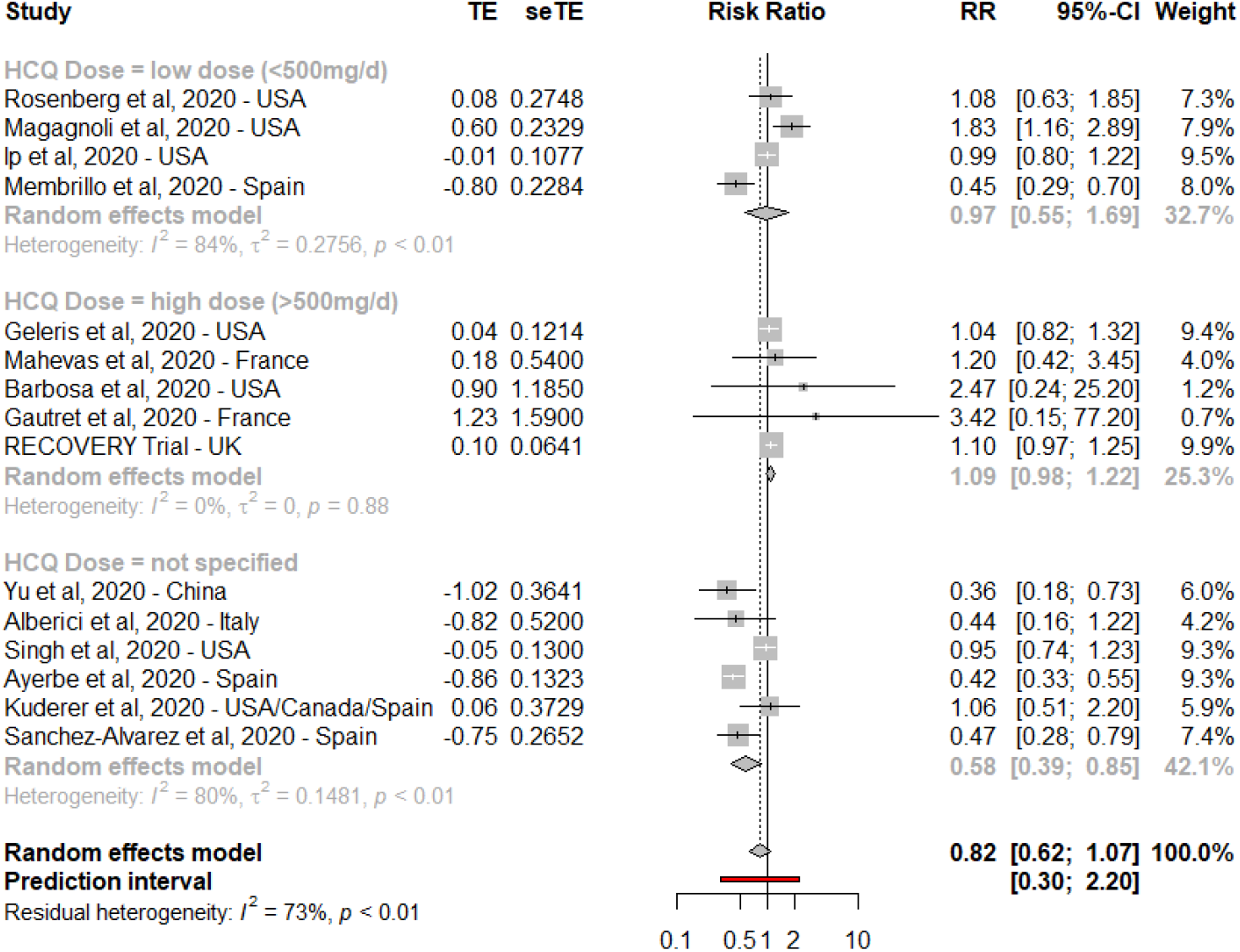
Forest plot for hydroxychloroquine alone and COVID-19 mortality risk, subgroup analysis per hydroxychloroquine dose

**Figure S7:**
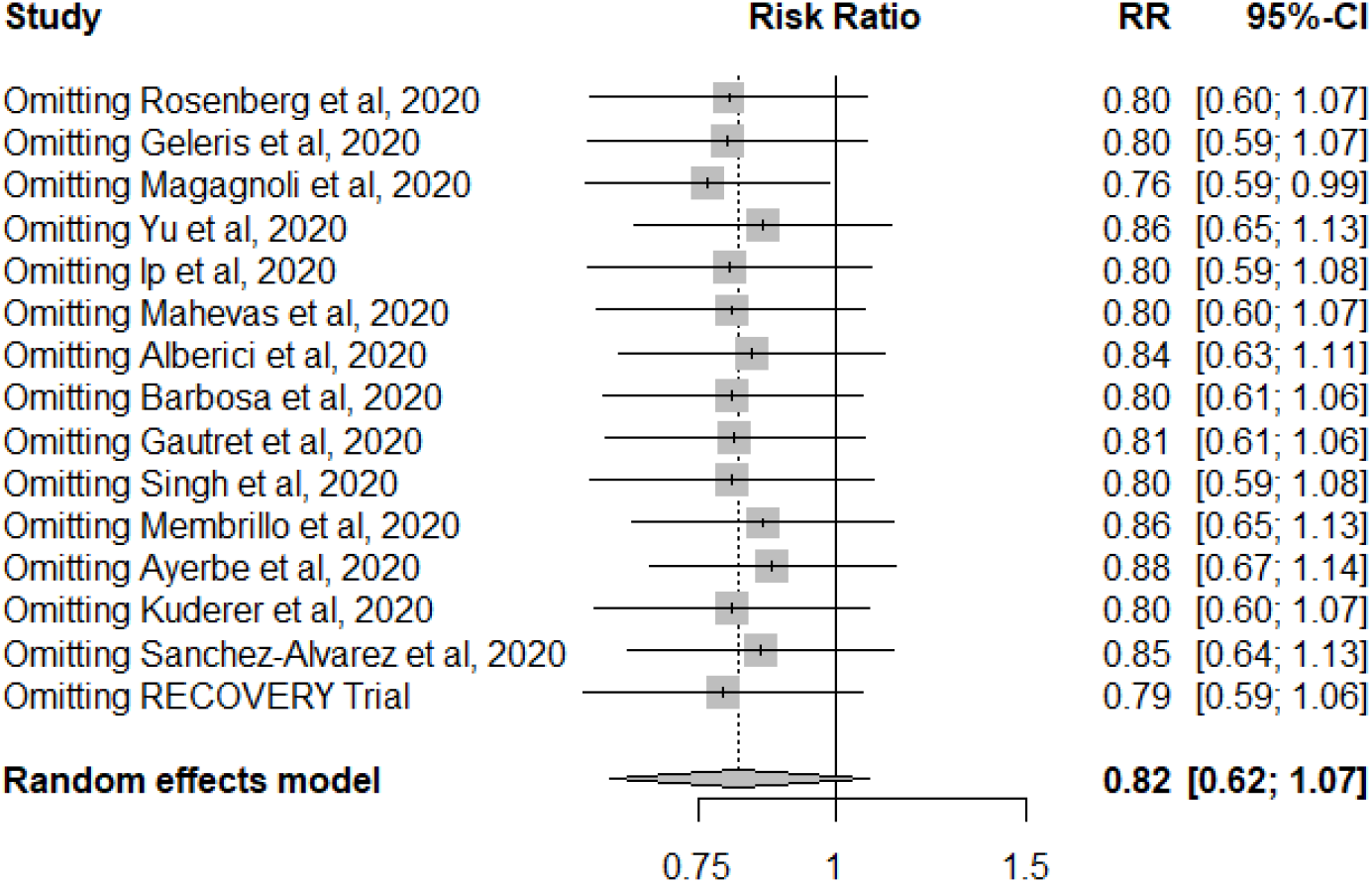
Influence analysis for hydroxychloroquine and COVID-19 mortality

## S1. Full electronic search strategy

### Cochrane Library

Website: https://www.cochranelibrary.com/advanced-search Cochrane Review matching (Hydroxychloroquine or HCQ) in Title Abstract Keyword AND (mortality or death) in Title Abstract Keyword AND (COVID-19 or SRAS-CoV-2) in Title Abstract Keyword - (Word variations have been searched)

PubMed

Website: https://pmlegacy.ncbi.nlm.nih.gov/pubmed/?term=(hydroxychloroquine+or+HCQ)+AND+(COVID-19+OR+SARS-CoV-2+OR+coronavirus)+AND+(Mortality+OR+death) ((hydroxychloroquine or HCQ) AND (COVID-19 OR SARS-CoV-2 OR coronavirus) AND (Mortality OR death)

### Web of Science

Website: http://apps.webofknowledge.com.proxy.insermbiblio.inist.fr/Search.do?product=UA&SID=F6KgcWI7K6kjXJwhAoH&search_mode=GeneralSearch&prID=9a27b347-ecf8-4832-9206-db1bbd2cc9a8 You searched for: TOPIC: (covid-19 OR SRAS-CoV-2) AND TOPIC: (hydroxychloroquine or HCQ) AND TOPIC: (mortality or death)

### Manual additional searches

#### MedRxiv

https://www.medrxiv.org/

Search: Hydroxychloroquine COVID-19 mortality

#### Google scholar

https://scholar.google.com/scholar?hl=en&as_sdt=0%2C5&q=hydroxychloroquine+COVID-19&btnG=

Search: Hydroxychloroquine COVID-19 mortality

## S2. PRISMA Checklist

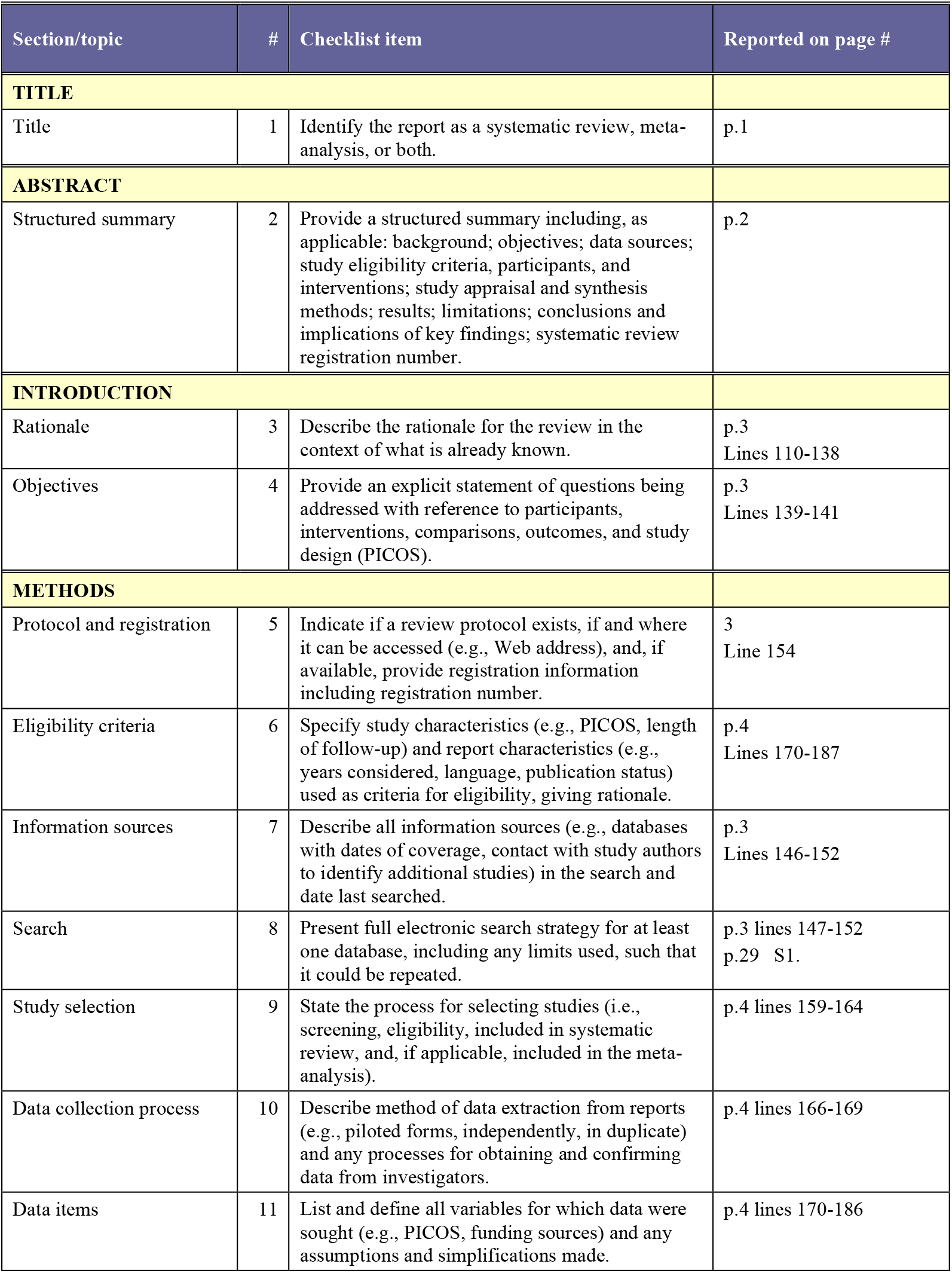
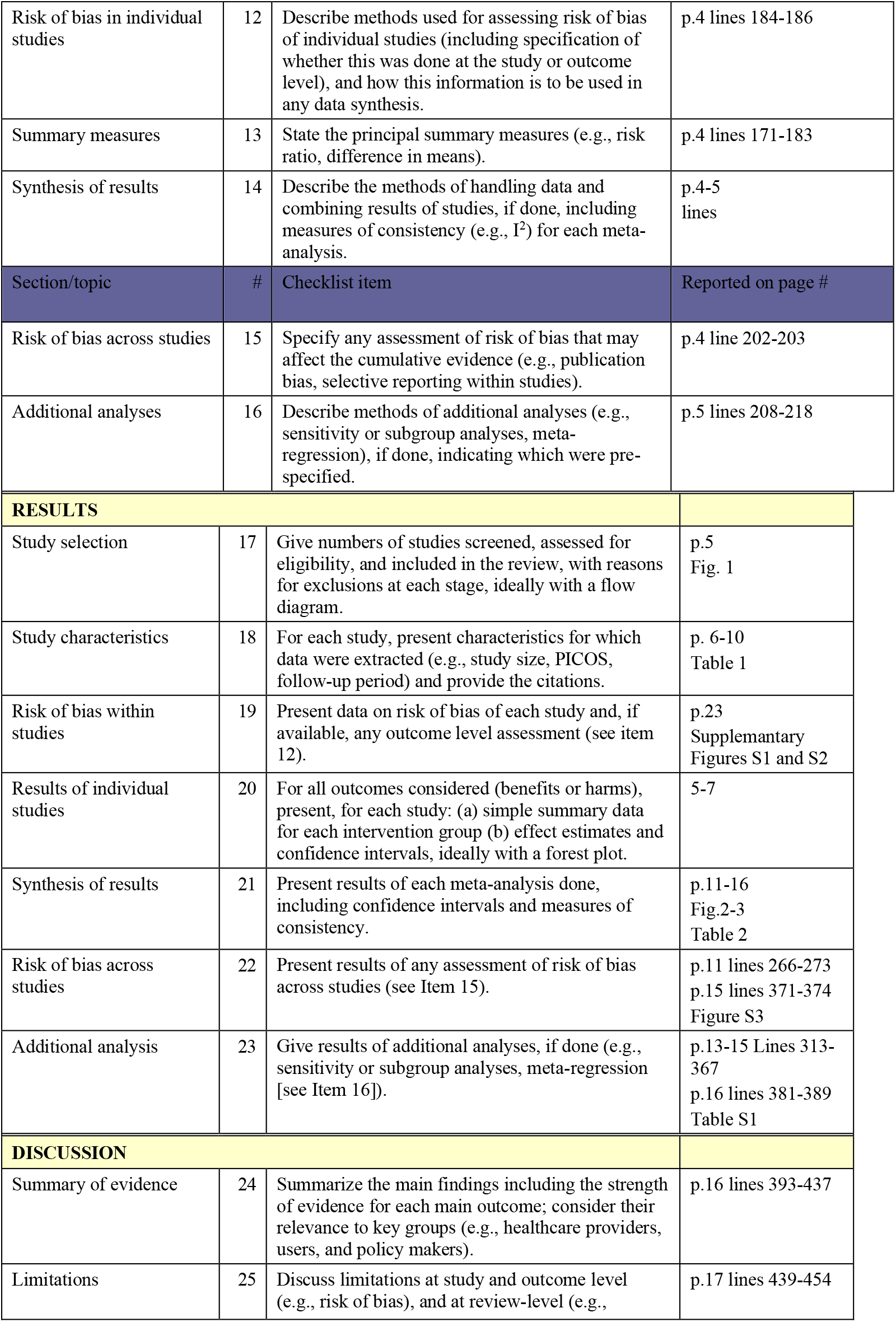
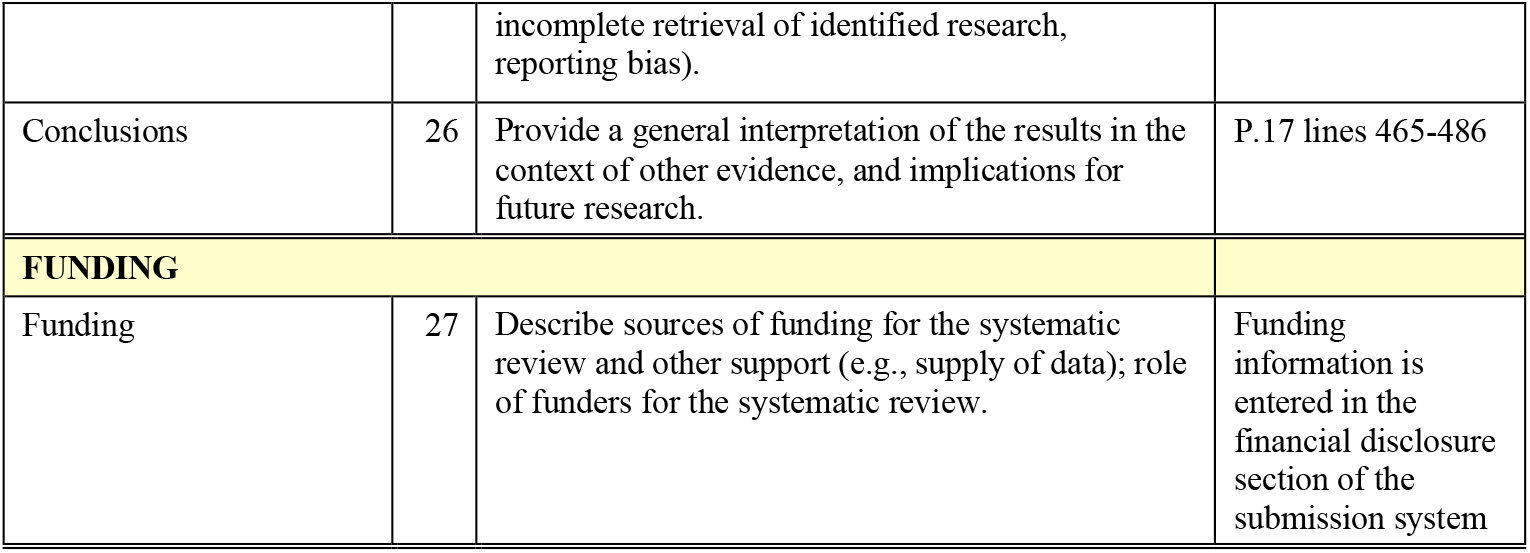

**Table S1:** Subgroup analysis for the associations between HCQ+AZI and mortality risk of patients with COVID-19 (observational studies)

| **Subgroup analysis for the associations between HCQ and mortality risk of patients with COVID-19 (excluding interventional studies)** |  |  |  |  |  |
| --- | --- | --- | --- | --- | --- |
|  | N | RRpooled | Heterogeneity |  |  |
|  |  |  | I<sup>2</sup> (%) | P<sub>within</sub> | P<sub>between</sub> |
| **HCQ alone** |  |  |  |  |  |
| All Studies | 13 |  |  |  |  |
| Type of article |  |  |  |  |  |
| Peer-reviewed | 9 | 0.76 [0.51-1.13] | 85% | <0.01 | 0.84 |
| Unpublished | 4 | 0.81 [0.52-1.27] | 72% | 0.01 |  |
| Adjusted estimate |  |  |  |  |  |
| Yes | 9 | 0.91 [0.67-1.24] | 70% | <0.01 | 0.0001 |
| No | 4 | 0.44 [0.35-0.55] | 0% | 0.52 |  |
| Risk estimated |  |  |  |  |  |
| Reported in the paper | 11 | 0.83 [0.61-1.11] | 72% | <0.01 | 0.82 |
| Calculated | 2 | 0.69 [0.15-3.25] | 54% | 0.14 |  |
| Risk of bias |  |  |  |  |  |
| Moderate | 5 | 1.02 [0.88-1.18] | 0% | 0.9 | 0.18 |
| Serious | 6 | 0.63 [0.38-1.04] | 89% | <0.01 |  |
| Critical | 2 | 0.75 [0.16-3.58] | 44% | 0.18 |  |
| Continents |  |  |  |  |  |
| America | 6 | 1.05 [0.93-1.19] | 30% | 0.2 | <0.0001 |
| Asia | 1 | 0.36 [0.18-0.73] | NA | NA |  |
| Europe | 5 | 0.45 [0.37-0.55] | 0% | 0.47 |  |
| Multiple | 1 | 1.06 [0.51-2.20] | NA | NA |  |
| Mean daily dose |  |  |  |  |  |
| Not specified | 6 | 0.58 [0.39-0.85] | 80% | <0.01 | 0.029 |
| <500 mg/d | 4 | 1.06 [0.84-1.33] | 0% | 0.75 |  |
| >500 mg/d | 3 | 0.58 [0.39-0.85] | 80% | <0.01 |  |
| Age |  |  |  |  |  |
| 63 years or less | 6 | 0.89 [0.64-1.24] | 59% | 0.03 | 0.39 |
| 64 years or more | 7 | 0.69 [0.43-1.10] | 89% | <0.01 |  |
| Cancer or hemodialysis patient based-population |  |  |  |  |  |
| No | 10 | 0.83 [0.59-1.18] | 85% | <0.01 | 0.34 |
| Yes | 3 | 0.61 [0.35-1.06] | 43% | 0.17 |  |
| Influence analysis (exclusion of Yu et al, Magagnoli et al, Membrillo et al, Ayerbe et al) | 9 | 0.95 [0.84-1.08] | 27% | 0.20 |  |

